# Environmental surveillance of poliovirus in four districts within two regions of Ghana

**DOI:** 10.1101/2022.07.02.22277180

**Authors:** Ernest Obese-Djomoah, Evangeline Obodai, Emmanuel Gberbi, Ewurabena Duker, Keren Attiku, Miriam Eshun, Bismarck Boahene, Samuel Victor Nuvor, John Kofi Odoom

**Affiliations:** Department of Microbiology and Immunology, School of Allied Health Sciences, University of Cape. Cape Coast, Ghana; Department of Virology, Noguchi Memorial Institute for Medical Research, University of Ghana, Legon, Ghana

**Author notes:** Corresponding author: John Kofi Odoom, Noguchi Memorial Institute for Medical Research, University of Ghana, P.O. Box LG 581, Legon, Accra, Ghana.

**Keywords:** Poliovirus, oral poliovirus vaccine, environmental surveillance, cytopathic effect, Ghana

## Abstract

The eradication of poliovirus is at its last phase through the efforts and strategies of Global Polio Eradication Initiation (GPEI). There are very few countries that are still endemic with wild poliovirus (WPV) and others with circulating vaccine derived poliovirus (cVDPV). The aim of the study was to detect silent circulation of WPV and VDPV in four districts within the Eastern and Volta region of Ghana. A systematic longitudinal design was used for the study. The convenient sampling technique was used to collect the samples every four weeks from two open and close sewage systems. The open sewage systems were located in New Juabeng and Ho districts while the close sewage systems were located in Asuogyaman and Ketu South districts. A total of 35 sewage samples were collected from September 2018 to May 2019. L20B and RD cell lines were used for the purification of poliovirus (PV) while real-time reverse transcriptase polymerase chain reaction (rRT-PCR) was use to characterize the serotypes of the PVs. The findings of the study showed that the prevalence of non-polio enterovirus (NPEV) and Sabin were 65.71% and 14.29% respectively. The characterized Sabins were serotype 1 and serotype 3 which were circulating in the two districts within the Eastern Region. The study did not detect any WPV and VDPV but isolated Sabin strains of the poliovirus. This necessitates the need for continuous environmental surveillance for poliovirus nationwide.

## Introduction

Poliovirus (PV) is a single-stranded positive-sense RNA virus which is a member of Human enterovirus C (HEV-C) and belongs to the family *Picornaviridae*, with the order of Picornavirales. It is the causative agent of poliomyelitis (commonly known as polio) which mainly affects children under the age of 5 years old. There are three serotypes of PVs: PV1, PV2 and PV3. Polioviruses are transmitted by the fecal-oral route; they multiply in the gut whether or not the infected persons are symptomatic [1].

The wild poliovirus type 2 (WPV2) was declared eradicated worldwide in September 2015 by the Global Commission for the Certification of Poliomyelitis Eradication (GCC). WPV2 was last detected in the year 1999 at Aligarh, Northern India, while the wild poliovirus type 3 (WPV3) has not been detected globally since November 2012. Africa Regional Certification Commission certified the WHO African Region as wild polio-free after four years without a case on 25^th^ August, 2020 [2]. AFP surveillance has been the gold standard for the detection of PV but paralytic poliomyelitis occurs in small factions of the individuals infected with the PV. This faction is further reduced as vaccination currently covers a large portion of the population globally, therefore rendering AFP surveillance insensitive to detect an outbreak that requires an immediate response [3].

Environmental surveillance (ES) serves as an additional method to monitor the transmission of poliovirus by testing sewage or wastewater samples, which may contain polioviruses in human faeces [4]. ES has been useful in detecting the presence of PVs circulating in districts whereby there has not been any detection of AFP from infected persons [4]. Combination of ES and AFP surveillances could be more sensitive to detect low circulation of WPV and circulating vaccine derived poliovirus (cVDPV), in order to sustain poliovirus eradication.

The last case of polio in Ghana was in November 2008, since then, all samples from AFP have tested negative for polio [5]. The sanitation in most parts of Ghana is poor, open defecation is widespread and surveillance indicators for AFP surveillance are not optimum in some regions. Nevertheless, the risk of importation of WPV from endemic countries such as Pakistan, Afghanistan and Nigeria is a real threat to Ghana as it has migrants and visitors from those countries. This study sort to detect silent circulation of wild type and vaccine-derived poliovirus in four districts within the Eastern and Volta Regions of Ghana.

## Method

### Selection of sample sites

Volta and Eastern regions were the selected regions purposely for the study. Ho is the capital of the Volta Region with an estimated population of about 1,649,523 inhabitants while Koforidua is the capital of the Eastern Region with an estimate population of about 2,917,039 inhabitants (estimated from the 2021 population census) [6].

There were three criteria that were taken into consideration for the selection of the two regions for the studies. (1) The districts were classified as high risk for PV transmission, based on existing data (i.e, poor surveillance indicators, high-risk population, sanitation, living conditions, routine immunization, and supplementary immunization activities coverage); (2) The sewer lines received waste from a considerable proportion of the population in the catchment area, with a minimum amount of waste coming from other areas; (3) There was absence of industrial waste in the proposed site.

### Sample collection, Transportation and Storage

Raw sewage and wastewater samples were collected from the selected sites in the morning between the hours of 8am to 10am. The running sewage and wastewater which effectively carries enteroviruses in human feaces were collected into 1.0 L plain plastic containers. The cup or bucket used for the collection was lowered gently into the sewage for the water to run into the container. The preferred spot for collection was where the velocity of the running waste water was high. The sewage was carefully transferred into plastic bottle using a funnel to avoid spillage on the bottle. closely tight and labelled with appropriate site identification names and epidemiological number. The samples were placed on a cold box with ice and sent immediately to the National Polio Reference Laboratory, Virology Department of the Noguchi Memorial Institute of Medical Research (NMIMR) where they were worked on the same day or kept at 4□C and processed the next day [6].

### Laboratory Procedures

#### Sewage Concentration

The sewage samples were taken out of the cold chain and allow to stand on the bench for 5 minutes for sedimentation of large solid material. A volume of 500 ml of the sewage sample were centrifuged for 20 min at 1500 g (minimum) at 4°C. The supernatants and the dry pellet were stored in 1 litre flask and 250ml centrifuge tube at 4°C respectively. The pH of the supernatant was adjusted to neutral (pH 7-7.4) using NaOH or HCl. If the supernatant was found to be acidic, approximately 0.2 ml 1N NaOH was added sequentially to it till the required pH was attained. A volume of 500 ml of the supernatant was poured into an Erlenmeyer flask for precipitation of the sewage. The rest of the corresponding raw waste water sample was kept at 4°C which served as backup until microscopy of the cell cultures inoculated with the concentrate shown absence of toxicity. After that it was stored at −20°C till all results were ready.

A concentration of 0.07M PEG6000, 0.56M dextran and 5.85M NaCl was added to 500ml of the supernatant. It was mixed thoroughly and kept in a constant agitation for 1 hour at room temperature using a magnetic stir plate at a speed sufficient to form a vortex. This continuous agitation was done at 4°C. One litre sterile conical separation funnel per sample was elevated and attached to a retort stand. Grease was spread on the gliding glass surfaces of the valves. Water tightness with a small volume of sterile water was checked. The valves were also checked to closed tightly. The mixture of the supernatant sewage sample with the reagents was poured into the separation funnels. The mixture was left to stand overnight at 4°C. The lower phase and the fuzzy interphase in the separation funnel were harvested carefully and slowly by drop-wise into a sterile tube. The dry pellet was resuspended from the centrifuge tube with a few millilitres of the harvested concentrate and added to the rest of the concentrate [6].

#### Treatment of Sewage Concentrate

Two millilitres of 20% chloroform stabilized with ethanol was added to 10ml of the suspension sample, 1 to 6g of sterile glass beads was added and vortexed briefly. The tube was later shaken vigorously for 20 minutes with Heidolph tube shaker. The tube was centrifuged according to the WHO Polio Laboratory Manual faecal suspension procedure (1500 g at minimum for 20 minutes at 4°C). The supernatant phase in the sterile 50ml centrifuge tube was aliquoted while the interphase and the deposits were discarded [6].

Penicillin G concentration of 100 IU/ml and 50µg/ml of gentamycin were added to the sample before inoculated on the cell culture medium. Two to four millilitres (ml) of the extracted concentrate was aliquoted and frozen at −20°C till the final results were determined. An approximate volume of 0.5 ml was inoculated on to the fresh monolayer cultures of L20B and RD cell lines. Five T-25 (25 cm^2^) flasks contained L20B cell lines while one T-25 flask contained RD cell lines per sample concentrate. The cultures in the flask were observed and marked for 5 days using the Poliovirus Investigation Worksheet [6].

#### Virus isolation

Virus growth was monitored daily by microscopy for cytopathic effect (CPE) for 5 days using the Poliovirus Investigation Worksheet. Samples that did not show CPE after 5 days were frozen and thawed twice, vortexed and re-inoculated and observed for CPE for another 5 days. Virus isolates were harvested after attaining 80-85% CPE and the infected cells frozen at −20°C. Virus isolate showing CPE on L20B cells were subjected to intratypic differentiation (ITD) using real-time reverse transcriptase polymerase chain reaction (rRT-PCR PCR) kit version 5.0 (provided by Centers for Disease Control and Prevention, Atlanta, USA). The ITD for poliovirus determined the serotype and the origin of the poliovirus while virus isolated on RD cells only were considered as non-polio enteroviruses [7].

### Identification and characterization

#### Poliovirus Real Time Reverse Transcriptase Polymerase Chain Reactions (Polio rRT-PCR ITD 5.0 Kit)

The rRT-PCR worksheet with name, date, primers, samples and sample order were filled out. Wells were named using thermocycler software for samples and controls. The Positive control constituted non-infectious control RNA supplied with Polio rRT-PCR kit while the reagent control constituted Quanta ToughMix + primer with no template. 1.5 ml Eppendorf tubes were labelled with primer names for each primer target. Virus isolated were thawed. PCR reagents were placed on ice and centrifuged. For each of the primers, 10µl Quanta ToughMix reagent was dispensed into each tube in the biosafety cabinet (BSC) designated for Master mix. Approximated volume of 8 µl nuclease free water was added. A volume of 1µ primer was added to the appropriate labelled eppendorf tube. A volume of 19µl of Master mix was dispense into appropriate PCR tube strips. A volume of 1µl of samples (cell culture supernatant) and controls were added to the appropriate PCR tube strips in the designated BSC. The PCR tube strips were centrifuged briefly at 5000 rpm and placed in rRT-PCR ABI 7500 machine. rRT-PCR conditions version 5.0; 40 cycles in each run: 50°C for 30 min; 95°C for 1 min; 95°C for 15 seconds; 50°C for 45 seconds; 25% ramp; 72°C for 5 seconds. Highlight annealing step was performed to collect product

#### Real time Reverse Transcriptase Polymerase Chain Reactions (Polio rRT-PCR VDPV 5.0 Kit)

The rRT-PCR worksheet with name, date, primers, samples and sample order as filled out. Wells were named using thermocycler software for samples and controls. The positive control constituted non-infectious control RNA supplied with Polio VDPV rRT-PCR kit and the reagent control constituted Quanta ToughMix + primer/probe with no template. The Eppendorf tubes (1.5 ml) for each primer target was labelled. The suspected virus isolates and PCR reagents were thawed on ice and centrifuged at 5000 rpm. For each primer set, 10µl Quanta ToughMix reagent was dispensed into each tube in the Master mix in the designated BSC. An approximate volume of 8µl nuclease free water was added. One microlitre (1µl) of primer/probe was added to the appropriate tube. Nineteen microlitres (19µl) of Master mix was added into the appropriate PCR tube strips. A volume of 1 µl of sample (cell culture supernatant) or control were added to the appropriate tubes in the designated BSC. The tubes were centrifuged briefly and placed in rRT-PCR ABI 7500. rRT-PCR conditions for version 5.0; 40 cycles in each run: 50°C for 30 mins; 95°C for 1 min; 95°C for 15 seconds; 50°C for 45 seconds 25% ramp; 72°C for 5 seconds. Highlight annealing step was performed to collect product.

#### Data Analysis

The laboratory data was compiled in MS excel which were imported into SPSS version 17 and analyzed. Univariable analysis of field investigation forms and isolations of viruses in the districts.

#### Ethical Consideration

The study ethical approval was obtained from the Scientific and Technical Committee of the Noguchi Memorial Institute, University of Ghana. The approval number was STC paper 3 (3) 2018-19.

## Results

### General characteristics and demography of study

A total of 36 sewage water samples from four sewage sampling sites situated in four districts (New Juabeng, Asuogyaman, Ho and Ketu South districts) within two regions were collected from September 2018 to May 2019 for the study. Some of the sewage water samples appeared cloudy while others appeared clear with suspended organic particles. None of the sewage samples were turbid or dark. The sewage systems that were reported in the study were open sewage system (50%) and close sewage system (50%). In each of the regions, at least there was a closed sewage system and an open sewage system where sampling took place as shown in table 1.

**Table 1:**
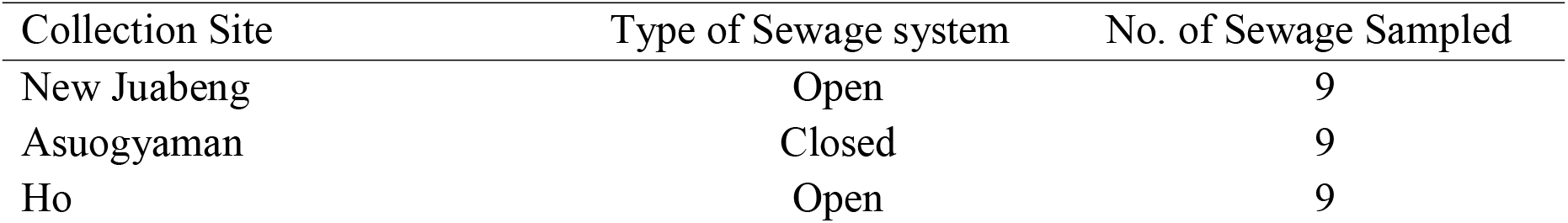
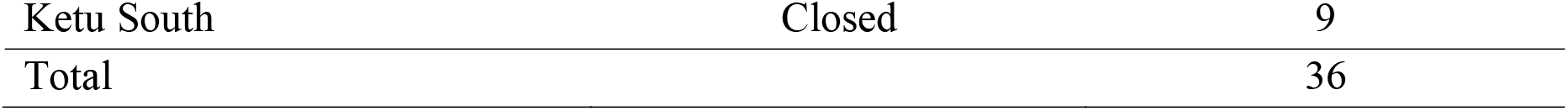
Sewage samples from collection sites

### Isolation from Cell Culture

Sewage samples collected from the Eastern region were inoculated in two cell lines; RD and L20B cell lines and observed for the first five days for cytopathic effect (CPE). It was observed that most of the monolayer cell cultures from the concentrated sewage samples showed CPE within 3-8 days’ incubation. It was observed that all the sewage concentrates from the New Juabeng collection sites showed CPE in the RD cell lines forming 25% of the total number of sewage that showed CPE in RD cell lines in the Eastern region. Sewage samples from Asuogyaman collection sites showed that most of the concentrate inoculated in the RD cell lines showed CPE (7, 19.44%) and only two samples never show CPE in the RD cell lines as shown in table 2.

**Table 2:**
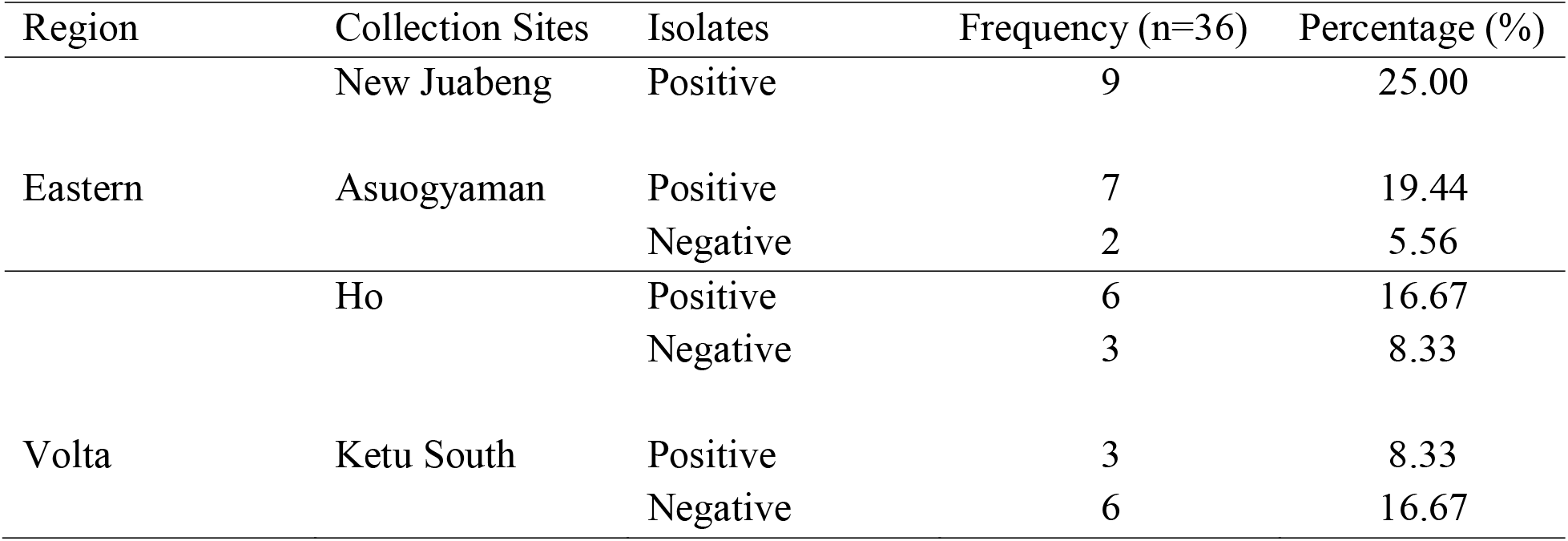
Collection Sites with virus isolates from Eastern and Volta Regions using RD cell line

There were also CPE in RD cell lines from sewages that were sampled from collection sites within the Volta region. It was observed that Ho sewage concentrates showed twice CPE in the RD cell lines (16.67%) as compared with the sewage samples collected from Ketu South (8.33%). It was also observed that most of the sewage from Ketu South did not show CPE in the RD cell lines (16.67%) as compared with the sewages concentrates from the Ho (8.33%). CPE in L20B cell lines showed more specific to PV as compared with other enteroviruses. The number of L20B cell lines that showed CPE for the concentrated sewage samples from the two collection sites (19.44%) were less than those that could not show CPE in the L20B (30.56%) for the two district sites under the Eastern region. The negative CPE in the L20B cell line was almost twice the sewage that showed CPE in the L20B cell lines. Sewage concentrates from the New Juabeng showed approximately twice of CPE in L20B (13.89%) more than the CPE in L20B from Asuogyaman sewage concentrates (5.56%) as shown table 3.

**Table 3:**
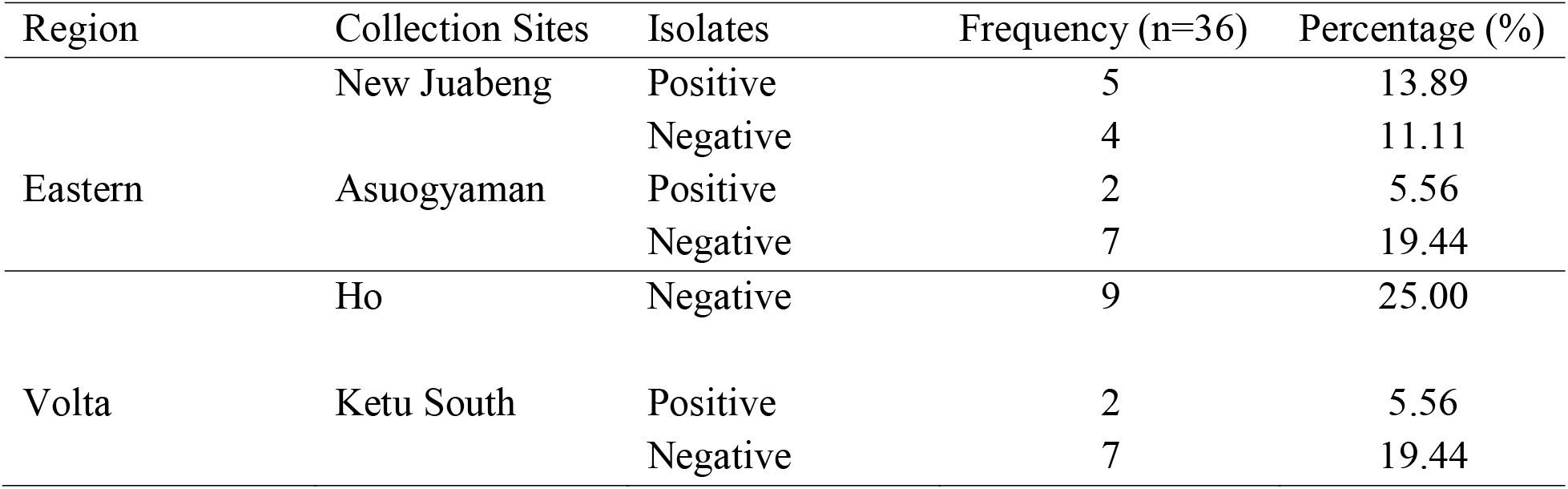
Collection sites with viral isolation from Eastern and Volta Regions using L20B cell line

## Discussions

### Processing of Sewage sample

Circulation of PV still remain a significant public health problem worldwide. The implementation of ES in countries that have not recorded WPV and VDPV over a long period through the AFP surveillance has become crucial. This is to monitor the importation of PV and immunization coverage in a catchment habitats or districts.

The current study applied the two-phase aqueous separation method as recommended by WHO in processing raw sewage samples. This processing aided to increase the virus concentration in the raw sewage which is free from other bacterial and fungal contaminations. This process was applied in ES studies conducted in Ghana [4], Malaysia [9], Senegal [10], Cameroon [11] and Nigeria [12] to determine viruses that might be circulating in the environment and likely to cause disease. An environmental surveillance studied in Moscow, Russian Federation, applied trap sampling method and 3% beef extract in 0.05 M Tris-HCl at a pH of 9.1 was used to process the raw sewage [13] while another studied at United Kingdom applied filtration and centrifugation methods using Centiprep YM-centrifugal concentration devices (Merck) [14]. Although WHO recommended two-phase separation method, the filtration and centrifugation methods are less laborious and quicker.

Almost all the sewage samples collected from the sample sites were not turbid which might be the outcome of few volume of the harvested pellets used for treatment and isolation of the PV and NPEV. The combination of the three compounds, PEG 6000, dextran T40 and NaCl might have facilitated the precipitation of the organic materials in the sewage samples to form the pellet harvest during antibiotic treatment of sewage samples and isolation of viruses as compared with WHO [8] manual that recommended two of the compounds, either PEG 6000 and Dextran T40 or NaCl.

### Virus Isolation

The detection of human enterovirus HEV confirmed the efficacy of the methods used to isolate the PV. The isolation rate of HEV was 71.43% which was consisted with studies that was carried out in Dakar, Senegal, 79.7% [15], as well as in Tehran province in Iran, 60.32% [16]. The HEV was relatively higher and was similar to the findings by Odoom et al., [4] in Ghana and in Fars province in Iran [16].

The isolation of HEV from site to site bases on the concentration of the sewage samples in the various sites were similar and are likely to be coxackiviruses, echoviruses, rhinoviruses and polioviruses. These HEV may be etiological agents associated in hemorrhagic conjunctivitis (AHC) also known as Apollo in Ghana. AFP and fulminant hepatitis may be caused by echoviruses whereas herpangina, Hand foot and Mouth disease may also be caused by coxsackieviruses. There are other echoviruses that may cause mild gastrointestinal and respiratory illness, myocarditis and encephalitis in children under 5 years of age. In the temperate climates HEV infection is common in the summer but the detection rate in this current studies shows that the seasonal pattern of detecting HEV is less evident in tropical areas where circulation trends are year round. The HEV might not necessarily be coming from the children under 5 years but adults within the habitat who might be discharging these viruses in the environment that will mix up with the children.

Cell culture is the gold standard for the isolation of PV and the study used two cell lines in isolation as recommended by WHO. Cytopathic effect (CPE) on the L20B cell lines should have been replicated on the RD cell lines within 3 to 5 days’ incubation. These shows the specificity in the isolation processes to ensure the specific type of viral isolation. Nevertheless, it was not all the CPE on the RD cell line was able to replicate on the L20B cell line. This also support the fact that no viruses are missed during isolation processes.

The isolation frequency of NPEV (3 isolates per month) was detected throughout the study period but there was a two-fold increment in January 2019. The detection of the NPEV in sewage was higher than detection of PV and it is consistent in work done on AFP surveillance on stools [13]. There were other studies with even higher rate of NPEV detection in Rio de Janeiro, Brazil [17] and Malaysia [9] from urban sewage. The sensitivity of the procedure applied in collecting the sewage samples and processing them to be able to detect NPEV indicates that the ES is adequately been performed. According to WHO, ≥ 30% of NPEV is expected to be isolated in samples processed by Grab method [8].

Five (17.86%) out of the nine (26.47%) suspected poliovirus from the L20B cell lines to the RD cell line arm was confirmed as Sabin-like isolate using rRTPCR. These findings were consistent with ES of PV carried out in Malaysia (10.43%) after 5 years of withdrawal from OPV to IPV [9]. Also similar to the studies done in Cameroon (7.2%) where PV was detected and characterized from urban sewages [11] and Nigeria Sabin detection rate of 29% [18].

There were isolations of PV in the Eastern region samples but none in the Volta region samples after the cell culture. This could be as a result of the modernization of properly disposing off children under 2 years’ stools which in turn could reduce the concentration of the virus if present in the large volume of waste that flows in the drainage system of the community. The habitat in the Volta region mostly use pit latrine to dispose off their human excreta while the few ones that use water closet would have their sewage water passing through the drainage system [19]. This might reduce the concentration of the PV and NPEV if they are circulating within the community. There is also the possibility of low immunization coverage among the children under 5 years so the vaccine strain was also not detected in the environment. The increase in temperature of the environment during the time of sampling was higher than room temperature (25□C) which might have been the effect of low survival of the virus in the sewage samples.

### Characterization of PV

The main target of ES is to detect WPV and circulating VDPV from raw sewage in a community but neither of these PVs were isolated from all the sewage collected. This was in consistent with findings in Malaysia [9], Senegal [10], Ghana [10] and Poland [20]. Probably, high immunization coverage in the selected districts and provinces may have contributed to the low isolation rates. Therefore, almost everyone in the population was protected from the WPV and VDPV infection. On the contrary, Nigeria [21], Pakistan [22] and Afghanistan [23] detected WPV and cVDPV in their environments. The findings indicated that wild polioviruses are still circulating among children in these nations.

The suspected PV after cell culture isolations were subjected to rRT-PCR for ITD to be able to determine the serotypes of the PV. The 5 isolates whose serotypes were determined came out to be Sabin like strain 1 and 3. It was determined that the most frequently detected serotype was PV3 which accounted for 66.67% of the total serotype strains. This finding is in consistent with similar findings from research works that have been carried out in Tehran [24] Japan [14] and Poland [20].

The presence of at least 1% of divergence in VP1 gene for serotypes 1 and 3 and 0.6% for serotype 2, in comparison with the prototype strains, classifieds them as VDPV. PVs with lower divergences than these are considered as “Sabinlike” [17]. This demarcation of ∼1% difference in nucleotide sequence of VP1 indicates that replication of the vaccine virus has occurred for approximately one year [25]. Isolates from the sewage samples indicates that there were no diversions in the VP1 gene.

## Conclusion

The hypothesis: there is no silent circulation of wild and vaccine-derived poliovirus in the selected districts of the Eastern and Volta regions of Ghana was supported. The ES detected circulation of Sabin 1 and Sabin 3 because of the bOPV that has been administered to children who are continually shedding the vaccine strain into the environment through their feaces. There was no Sabin isolation in the Volta region. There is also the possibility of low immunization coverage among the children under 5 years so the vaccine strain was also not detected in the environment. ES has proven to be very sensitive and it should be implemented in all the districts thought out the whole country. This will facilitate the early detection of any importation of the PV and reemergence strain of the PV as the world put forces together to eradicate polio.

## Data Availability

All data produced in the present work are contained in the manuscript

## Authors’ contributions

Conceptualization and design: EOD, JKO, EO. Data collection and Statistical analysis: EOD, ME, KA, EG, ED, BB. Drafting of the manuscript, integrity of the data, and the accuracy of the data analysis: EOD, JKO, EO, SVN. All authors have read and agreed to the final manuscript.

## Consent for publication

Not applicable

## Ethics approval and consent to participate

Not applicable

## References

1. Pallansch, M. A., Oberste M. S. Wjl. Enteroviruses: Polioviruses, Coxsackieviruses, Echoviruses, and Newer Enteroviruses. In: Fields Virology. Knipe D. M., Howley P. M.,. Philadelphia, PA Lippincott, William Wilkins. 2013; Eds. (6th):490–526.

2. World Health Organization. Global polio eradication initiative applauds WHO African region for wild polio-free certification. WHO int news [Internet]. 2020; Available from: https://www.who.int/news/item/25-08-2020-global-polio-eradication-initiative-applauds-who-african-region-for-wild-polio-free-certification

3. Nathanson, N. & Kew OM. From emergence to eradication: the epidemiology of poliomyelitis deconstructed. Am J Epidemiol. 2010;172:1213–1229.

4. Odoom J., Obodai E, Diamenu S, Ahove V, Addo J, Banahene B, et al. Environmental Surveillance for Poliovirus in Greater Accra and Eastern Regions of Ghana-2016 Virology□: Current research. 2017;1(1):1–6.

5. Odoom JK, Afia N, Ntim A, Sarkodie B, Addo J, Minta-asare K, et al. Evaluation of AFP surveillance indicators in. 2014;14(1):1–8.

6. Ghana Statistical Service. 2021 Population and Housing Census Press Release on Provisional Results. Press Release [Internet]. 2021;(September):1–7. Available from: www.census2021.statsghana.gov.gh

7. Jk O, Obodai E, Diamenu S, Ahove V, Addo J, Banahene B, et al. Environmental Surveillance for Poliovirus in Greater Accra and Eastern Regions of Ghana-2016 Virology□: Current research. 2017;1(1):1–6.

8. WHO. Manual for the virological investigation of poliomyelitis. 2004;

9. Yusof MA, Jahis R, Judson J, Ismail AK, Zauri M, Wahid A, et al. Clinical Biotechnology and Microbiology Environmental Surveillance of Polioviruses□: Five Years ‘ Experience in Malaysia Urban Population Before and After Withdrawal of Oral Polio Vaccine. 2018;2(4):392–400.

10. Ndiaye, A. K., Mbathio, P. A. & Diop OM. Environmental surveillance of poliovirus and non-polio enterovirus in urban sewage in Dakar, Senegal (2007–2013). Pan Afr Med J. 2014;19:1–7.

11. Njile DK, Alain S, Mba S, Claire M, Zanga E, Mengouo MN, et al. Detection and characterization of polioviruses originating from urban sewage in Yaounde and Douala, Cameroon 2016 – 2017. BMC Res Notes [Internet]. 2019; Available from: https://doi.org/10.1186/s13104-019-4280-6

12. Adekunle Adeniji J, Oladapo Adewale A, Cephas Faleye TO, Olubusuyi Adewumi M. Isolation and Identification of Enteroviruses from Sewage and Sewage-Contaminated Water Samples from Ibadan, Nigeria, 2012-2013. J Virol Antivir Res. 2017;06(03):2012–3.

13. Ivanova OE, Yarmolskaya MS, Eremeeva TP, Babkina GM, Baykova OY, Akhmadishina L V, et al. Environmental Surveillance for Poliovirus and Other Enteroviruses: Long-Term Experience in Moscow, Russian Federation, 2004–2017. Virusus MDPI. 2019;1–13.

14. Majumdar M, Klapsa D, Wilton T, Akello J, Anscombe C, Allen D, et al. Isolation of Vaccine-Like Poliovirus Strains in Sewage Samples From the United Kingdom. 2018;217.

15. WHO. Guidelines for environmental surveillance of poliovirus circulation. World Heal Organ Geneva Dep Vaccines Biol WHO. 2003;

16. Ndiaye, A. K., Mbathio, P. A. & Diop OM. Environmental surveillance of poliovirus and non-polio enterovirus in urban sewage in Dakar, Senegal (2007–2013). Pan Afr Med J. 2014;19:1–7.

17. Momou KJ, Akoua-Koffi C, Akre DS, Adjogoua EV, Tiéoulou L et al. Detection of enteroviruses in urban wastewater in Yopougon, Abidjan. Pathol Biol. 2012;60:e21–6.

18. Jr IPS, Burlandy FM, Oliveira SS, Amanda M, Sousa C, Silva EM, et al. Acute flaccid paralysis laboratorial surveillance in a polio-free country□: Brazil, 2005 – 2014. Hum Vaccin Immunother [Internet]. 2017;13(3):717–23. Available from: http://dx.doi.org/10.1080/21645515.2016.1236164

19. Weldegebriel, G.; Adeneji, A.; Gasasiral, A.; Okello, D.; Elemuwa C. et al. Environmental Surveillance for Poliovirus in Polio High Risk States of Nigeria, 2011-2012. Sci J Pub Hlth. 2015;3:655–63.

20. Nakamura T, Hamasaki M, Yoshitomi H, Ishibashi T, Yoshiyama C, Maeda E, et al. Environmental Surveillance of Poliovirus in Sewage Water around the Introduction Period for Inactivated Polio Vaccine in Japan. Appl Environ Microbiol. 2015;81(5):1859–64.

21. Figas A, Wieczorek M, Żuk-wasek A. Isolation of Sabin-like Polioviruses from Sewage in Poland. 2018;67(1):89–96.

22. Gumede N, Okeibunor J, Diop O, Baba M, Barnor J, Mbaye S, et al. Progress on the implementation of environmental surveillance in the African Region, 2011-2016. J Immunol Sci. 2018;2(SI1):24–30.

23. Moran-Gilad, J.; Kaliner, E.; Gdalevich, M.; Grotto I. Public health response to the silent reintroduction of wild poliovirus to Israel. J Clin Microbiol Infec. 2016;22:140–S145.

24. Kroiss SJ, Ahmadzai M, Ahmed J, Alam MM, Chabot-couture G, Id MF, et al. Assessing the sensitivity of the polio environmental surveillance system. 2018;1–18.

25. Matrajt G, Naughton B, Bandyopadhyay AS, Meschke JS. A Review of the Most Commonly Used Methods for Sample Collection in Environmental Surveillance of Poliovirus. 2018;67(Suppl 1).

26. Noori N, Drake JM, Rohani P. Comparative epidemiology of poliovirus transmission. Sci Rep [Internet]. 2017;1–12. Available from: http://dx.doi.org/10.1038/s41598-017-17749-5

